# Retrospective impact evaluation of indoor residual spraying in Côte d’Ivoire using routine malaria surveillance data: the role of contemporaneous covariates and data quality

**DOI:** 10.1101/2025.04.30.25326428

**Authors:** Christian Selinger, Kouakou D Appeti, Sumaiyya Thawer, Georgina Angoa, Serge A Aïmain, Serge B Assi, Emilie Pothin, Emily R Hilton, Antoine M Tanoh

**Affiliations:** Swiss Tropical and Public Health Institute, Allschwil, Switzerland; University of Basel, Basel, Switzerland; Programme National de Lutte Contre le Paludisme, Abidjan, Côte d’Ivoire; Institut Pierre Richet (IPR), Institut National de la Santé Publique (INSP), Bouaké, Côte d’Ivoire; Centre Suisse de Recherches Scientifiques en Côte d’Ivoire, Abidjan, Côte d’Ivoire; PMI VectorLink Project, PATH, Seattle, Washington, USA

## Abstract

**Objectives:** Indoor residual spraying (IRS) was introduced in Côte d’Ivoire in 2020. We seek to identify potential factors that could compromise the quality of routine malaria incidence and determine how impact quantification of IRS on malaria incidence depends on data quality by emphasizing robust counterfactuals with contemporaneous covariates.

**Methods:** Using mixed-effect models and correlation analysis we compare data from two sources (routine surveillance and re-digitized registries). To quantify decrease in incidence post intervention, we apply Bayesian structural time series analysis with posterior feature selection to determine impact confounders.

**Results:** The presence of IRS, test positivity and reporting rates are significant factors impacting data quality. For high-quality data, we estimate a 12% resp. 19% decrease in incidence rate one year after the intervention, while for low-quality data estimates are 30% resp. 42%. Important covariates for counterfactual modeling are control district incidence, reporting and test positivity rates.

**Conclusions:** Improvements in data quality by scrutinizing test positivity rates and health facility reporting can help avoid overly optimistic impact estimations for IRS. Incidence control districts remain inevitable in order to factor out impact confounders.

## Introduction

After two decades of steadily declining malaria mortality from 2000 to 2019, global progress in the fight against malaria has recently stalled and, in some regions, reversed (1). In Côte d’Ivoire, Malaria remains a leading cause of morbidity and mortality, accounting for around 33% of outpatient visits at health facilities (2). Vector control interventions including indoor residual spraying (IRS) and insecticide-treated nets (ITNs) remain important cornerstones of malaria prevention (3). Côte d’Ivoire included IRS in its National Malaria Strategic Plans 2016–2020 and 2021–2025 in order to reduce malaria burden in high transmission districts (2). IRS was introduced in two districts in 2020 and carried out annually until 2022. After 2022, IRS was stopped due to loss of funding through the US President’s Malaria Initiative (PMI), despite the evidence of its impact on clinically relevant outcomes in ‘real-world’ settings (4). Compared to randomized control trials (RCTs), real-world evidence from routine surveillance systems is less costly to produce, specific to the local context and of practical use to decision-makers. On the other hand, such analyses do not typically use any control groups or counterfactuals which is crucial to all retrospective impact evaluations (5–7). Counterfactuals should represent the temporal evolution of the outcome measure in the hypothetical absence of the intervention. Clinical impact of vector control can be estimated using confirmed malaria cases. Every month, such data are reported to a centralized database based upon facility-level paper registries using the District Health Information Software (DHIS2) system. The National Malaria Control Program (NMCP) of Côte d’Ivoire has implemented the DHIS2 database as early as 2015 with only recently improved consistency and validity (8). In a project aiming at assessing the impact of IRS, PMI in collaboration with the NMCP re-digitized key malaria indicators from health facility consultation registers during the years 2019-2022 of four districts: Nassian and Sakassou with IRS campaigns conducted in 2020&2021, and Dabakala and Beoumi as control districts. Based on this data (henceforth referred to as PMI data), a comprehensive impact evaluation estimated a significant reduction in clinical incidence following the IRS campaigns (9).

Complementary to the impact quantification undertaken in (9), our analysis has two main objectives. First, we seek to identify potential factors contributing to the differences between data reported into DHIS2 and the data re-digitized by PMI, assuming that DHIS2 data are generally of lower quality and compromised at the level of data entry and compilation.

Second, we utilize a model-based approach to interrogate how differences in data quality alter intervention impact quantification. We emphasize building counterfactuals incorporating a large number of contemporaneous covariates relying on Bayesian structural time series analysis. This technique has been suggested as a viable alternative to interrupted time series analysis for health impact assessment (10,11) with clinical, routine and remote-sensed data (12–14) and probabilistically combines covariates such as rainfall, vegetation cover, temperature, but also routine health surveillance quality and most importantly reported malaria incidence from control districts to factor out possible confounders of IRS impact.

The results of this analysis should help promote critical use of routine malaria surveillance data for decision-making. Intensified data validation and audits combined with analytics that allow identifying potential causal confounders should further add to the existing body of evidence on how IRS affects malaria incidence in the ‘real’ world. In light of increasingly limited resources for malaria surveillance, treatment and prevention, the importance of high quality and appropriately contextualized data for decision making to National Malaria Control Programs cannot be overestimated.

## Methods

### Data sources and quality assessment

We extracted and post-processed routine surveillance data for malaria from the DHIS2 database in collaboration with the NMCP of Côte d’Ivoire. In the country, the reporting follows the hierarchy of referral in the sanitary pyramid, i.e. health facilities in the periphery with recently added community health workers (CHW) and local general hospitals (Hôpital Général, HG) refer to regional hospitals (Centre Hospitalier Régional, CHR) who refer to university hospitals (Centre Hospitalier Universitaire, CHU). The nested spatial organization consists of 113 health districts within 26 health regions that report to the Ministry of Public Hygiene and Universal Health Coverage at the national level. We considered clinical malaria cases of all ages reported at the level of health districts. These monthly data were entered into DHIS2 from paper registries in health facilities, HG, and CHR by local health workers and validated by the NMCP. As for CHW and CHU, inconsistency in reporting across districts and time, prohibited further consideration for this analysis. Since presumed cases (i.e. without confirmed tests) have been reported into the DHIS2 database only since 2021, we considered the difference between suspected and tested cases as presumed cases whenever the latter were not directly available in the database.

In preparation of the impact evaluation for IRS in the districts Nassian and Sakassou with control districts Dabakala and Beoumi, PMI decided to collect data directly from health facility registries in the four study districts due to insufficient data quality (15). Patient data were abstracted from consultation registries (“registres de consultations curatives”) for the period September 2018 to April 2022, we refer to details of inclusion criteria to recently published study results (9).

In order to control for factors unrelated to the intervention, we also included environmental, entomological and surveillance covariates into the causal impact analysis (see Supplementary Table 1). Remote-sensed data (rainfall, average daily temperature, leaf cover, average daily humidity) was extracted from the ERA5 database (16), aggregated monthly at the health-district level. Entomological covariates (monthly indoor and outdoor biting rates and monthly entomological inoculation rates) were extracted from the PMI VectorLink annual entomological reports for the years 2019-2021 (17–19) for the four districts of interest. To take into account changes in the health surveillance we also derived from the DHIS2 database the following district-level monthly quantities: health facility reporting rate (short: hf_reporting), monthly testing rate (short: testing), test positivity rate (short: tpr_total), the number of active health facilities (short: N_hf_active) and coverage of intermittent preventative therapy for pregnant women for the first trimester (short: ipt1_coverage). Although the latter quantity concerns only a small proportion of the population at risk, we consider it as a general proxy for access to health care. We normalized and scaled all covariates prior to further analysis.

### Incidence rate calculations with adjustments

To adjust incidence (to both DHIS2 and PMI crude data), we used the following epidemiological indicators. First, we extracted monthly confirmed malaria cases reported and validated at the level of health districts in the DHIS2 system or by PMI respectively. Second, we used presumed cases (short: pr) from the DHIS2 database which are defined as cases with malaria-like symptoms without having been tested. Third, we calculated the test positivity rate (short: tpr) as the ratio of confirmed over tested cases. Here, we assumed that cases were tested with either blood spot or rapid diagnostic test (RDT). For the reporting rate, we considered for each health district the ratio of months within a year, where health facilities reported into the DHIS2 system. Finally, the health care seeking rate (short: s) was extracted from the 2016 Malaria Indicator Survey (20) and the 2021 Demographic and Health Survey (DHS) report (21) estimates of health care seeking for children with fever made at the regional level. Sakassou, Beoumi and Dabakala are located in the Center-North region and Nassian in the North East region. We interpolated these values linearly between the years.

We then calculated adjustments to incidence rates for both data sources DHIS2 and PMI using the same quantities as detailed in Supplementary Table 2. District-level population data were extracted from the 2021 census data with 3% annual growth rate extrapolation. We calculated incidence rate as the ratio of adjusted incidence over district population.

### Multiple imputation of missing data

In the period of interest, we have detected 6.7% of missing data among all contemporaneous covariates across the four health districts (see Supplementary Figure 1). We used the R package Amelia (22) to perform multiple imputations of missing data for a subset of contemporaneous variables (biting_rate_hcl_outdoor, biting_rate_hcl_indoor, EIR_ibpn), with health district as cross-sectional variable and the month during the year as temporal variable to account for seasonality. Based on the rule-of-thumb we performed 7 imputations for 7% of missing data. We averaged imputed data points and used them to replace missing data in the original data set. In addition, we also imputed two incidence points for the Beoumi district, which had suspicious outliers.

### Mixed effect models to analyze difference between data sources

In order to identify associations between difference in incidence rates recorded in the DHIS2 and PMI data sets, we performed a mixed-effect linear model analysis (23) on the difference of log-transformed incidence data with a random intercept for the health district. We used a backward stepwise model selection approach (24) to eliminate sequentially factors from two models (see Supplementary Tables 3&4): *model1* had only health system covariates (IRS intervention, hf_reporting_rate, N_hf_active, tpr_total, ipt1_coverage), *model2* had in addition also environmental variables (rainfall, temperature, leaf, humidity, biting rates, EIR) without and with one month lag obtained from cross-correlation analyses. For the resulting parsimonious model, we determined the coefficient estimators and two-sided p-values.

### Correlation analysis between incidence rates and contemporaneous variables

We performed a Pearson correlation analysis between monthly incidence rates and contemporaneous variables for lags of up to one month in the past. This analysis was performed across health districts and per incidence data source.

### Bayesian structural time series analysis for impact quantification

Bayesian structural times series analysis (25,26) is an extension of classical time series analysis techniques such as auto-regression moving average models. The method integrates elements of state-space models (i.e. observation and hidden state parameters are estimated concurrently) and Bayesian feature selection (e.g. through spike-and-slab priors) and provides forecasting of time series with high-dimensional time-dependent covariates. We utilize this method to construct counterfactuals for intervention impact quantification. For monthly outcome measures (malaria incidence rates or EIR) we implement local linear and seasonal effects, and use contemporaneous regressors for lagged time series that are independent from the incidence rates (e.g. rainfall, temperature). In order to train the pre-intervention model we use either the incidence data in the IRS districts (control: “none”), incidence data in both IRS and non-IRS districts (control: “incidence), or all contemporaneous covariates including incidence from all districts (control: “all covariates) but excluding entomological covariates, since those are supposed to be statistically dependent on incidence. Similarly, for the impact analysis with EIR, we use EIR data in IRS districts only (control: “none”), EIR from both IRS and non-IRS districts (control: “EIR”), or all contemporaneous covariates except incidence covariates, since those are supposed to be statistically dependent on EIR.

From the Bayesian feature selection for models with contemporaneous covariates, we extract marginalized posterior distribution of regression parameters to quantify how often a regressor has been included in the model, but also its magnitude and whether 95% of the probability mass was either positive or negative (“consistent regressors”). Causal impact is determined by the difference of predicted incidence in the absence of the intervention (i.e. from the counterfactual) and the actually recorded data in presence of the intervention, averaged over one-year after the intervention deployment. We consider causality as established if in less than 5% of 10000 simulated counterfactuals we observe incidence inferior to the observed incidence with IRS.

## Results

### Factors influencing the difference in incidence between DHIS2 and PMI data

After adjusting crude incidence of malaria cases from DHIS2 and PMI data sources (Table 2, adjustment 2), we observed a range between 15 and 100 monthly incidence points per 1000 population between September 2018 and April 2022 with peaks occurring generally between July and October (Figure 1). Two consecutive months for the control district of Sakassou had zero incidence recorded in the DHIS2 data. We considered these as missing data and imputed them using multiple imputation algorithms (see Methods). PMI incidence rates were generally higher than those based on DHIS2 data, and Sakassou followed more consistently the rainfall seasonality trends than Nassian. The impact analysis focused on incidence data until August 2021, date of the start of the second deployment of IRS. Analyzing the difference between incidence rates from the two data sources including model selection for a comprehensive set of contemporaneous and categorical variables (see Supplementary Table 2&3) suggested that there were several statistically significant factors explaining the difference. We identified the presence of the first IRS intervention, increased IPT1 coverage and test positivity rates to be statistically associated with a decrease in difference (P<0.05) Table 1.

**Figure 1:**
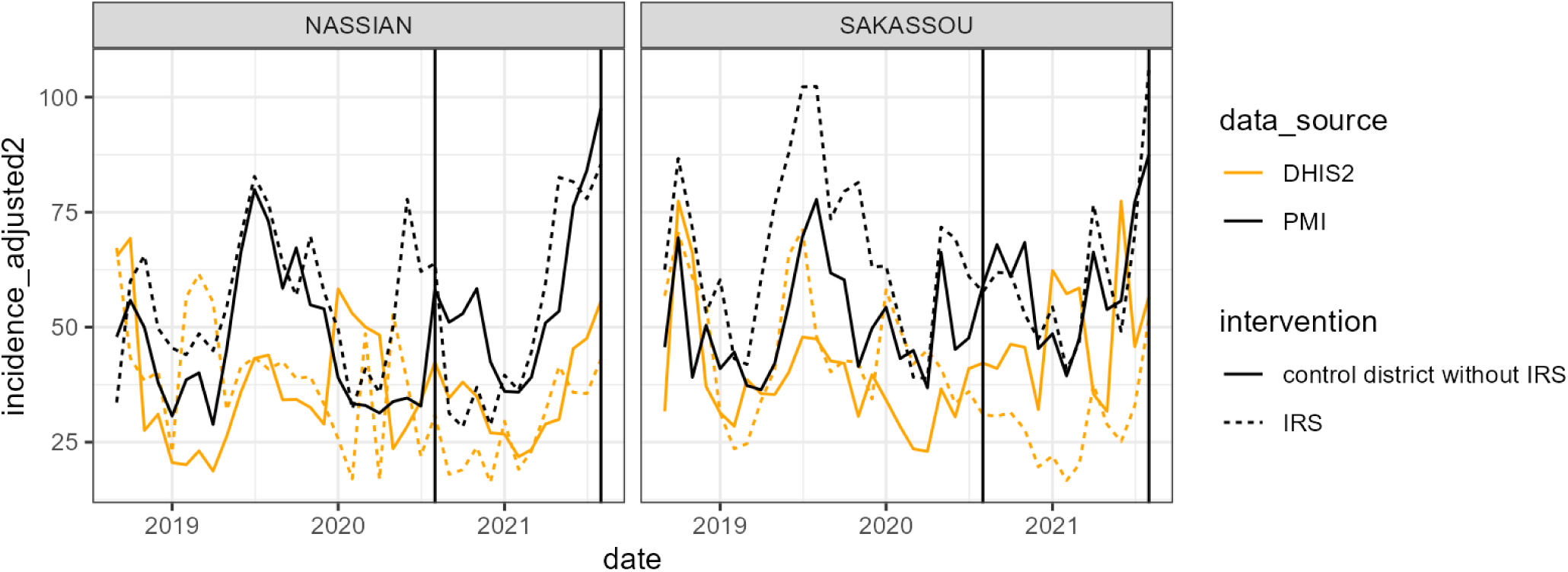
Adjusted monthly incidence rates of confirmed malaria cases in the IRS-treated districts of Nassian and Sakasso (solid lines) and their respective control district (dashed lines). The color code refers to the data source (DHIS2 in orange, PMI in black).

**Table 1:**
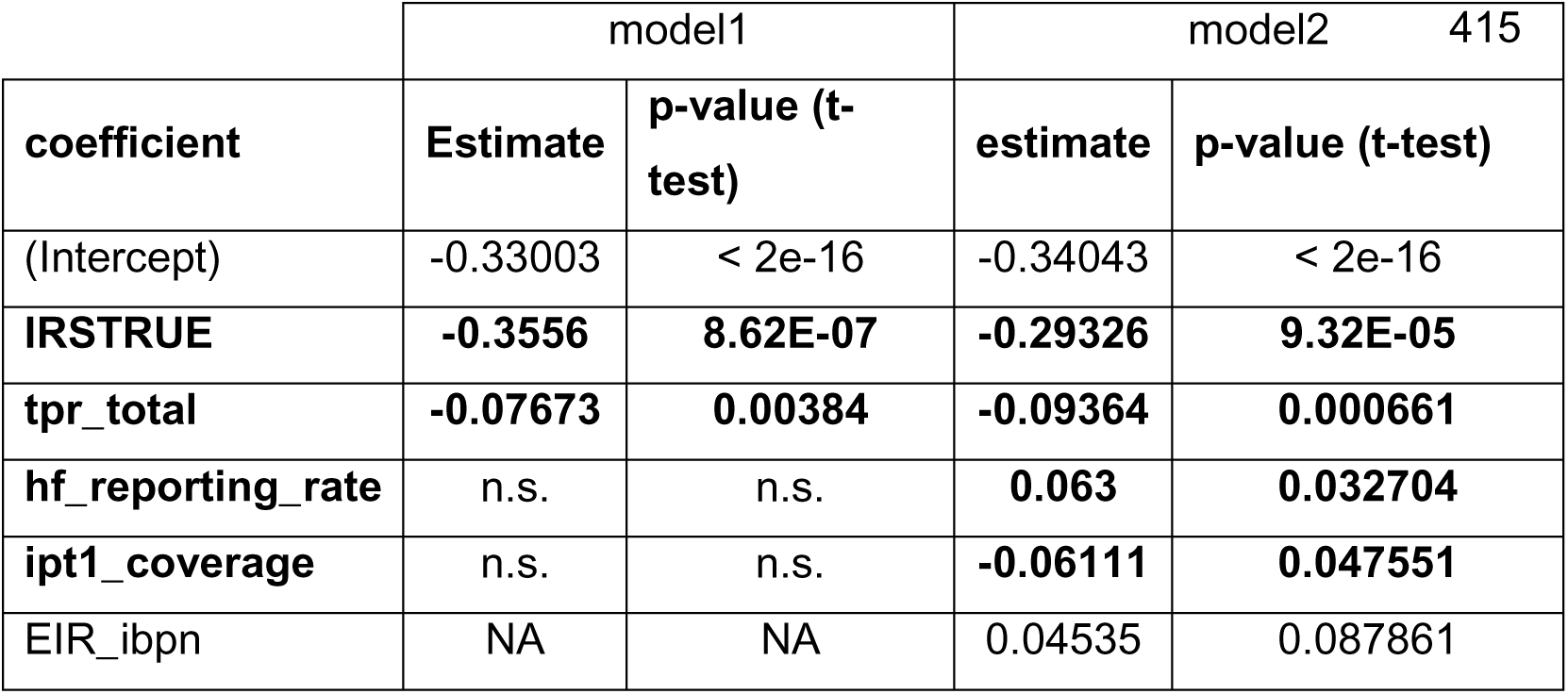
Results table of best log-linear mixed-effect model of incidence difference between DHIS2 and PMI data identified after model selection. *model1* considered only health system related covariates, whereas *model2* also included entomological and environmental covariates.

### Correlations between incidence and contemporaneous variables

For the correlation analysis between monthly incidence rates and contemporaneous variables, we considered both data from the same month and with a one-month lag across all four districts before and after the intervention (Figure 2). We noticed that entomological variables were highly correlated amongst each other (e.g. 0.57 and 0.53 Pearson correlation coefficient between EIR and indoor or outdoor biting rates). Biting rates also positively correlated with malaria incidence for the PMI data (0.53 and 0.57 correlation coefficient).

**Figure 2:**
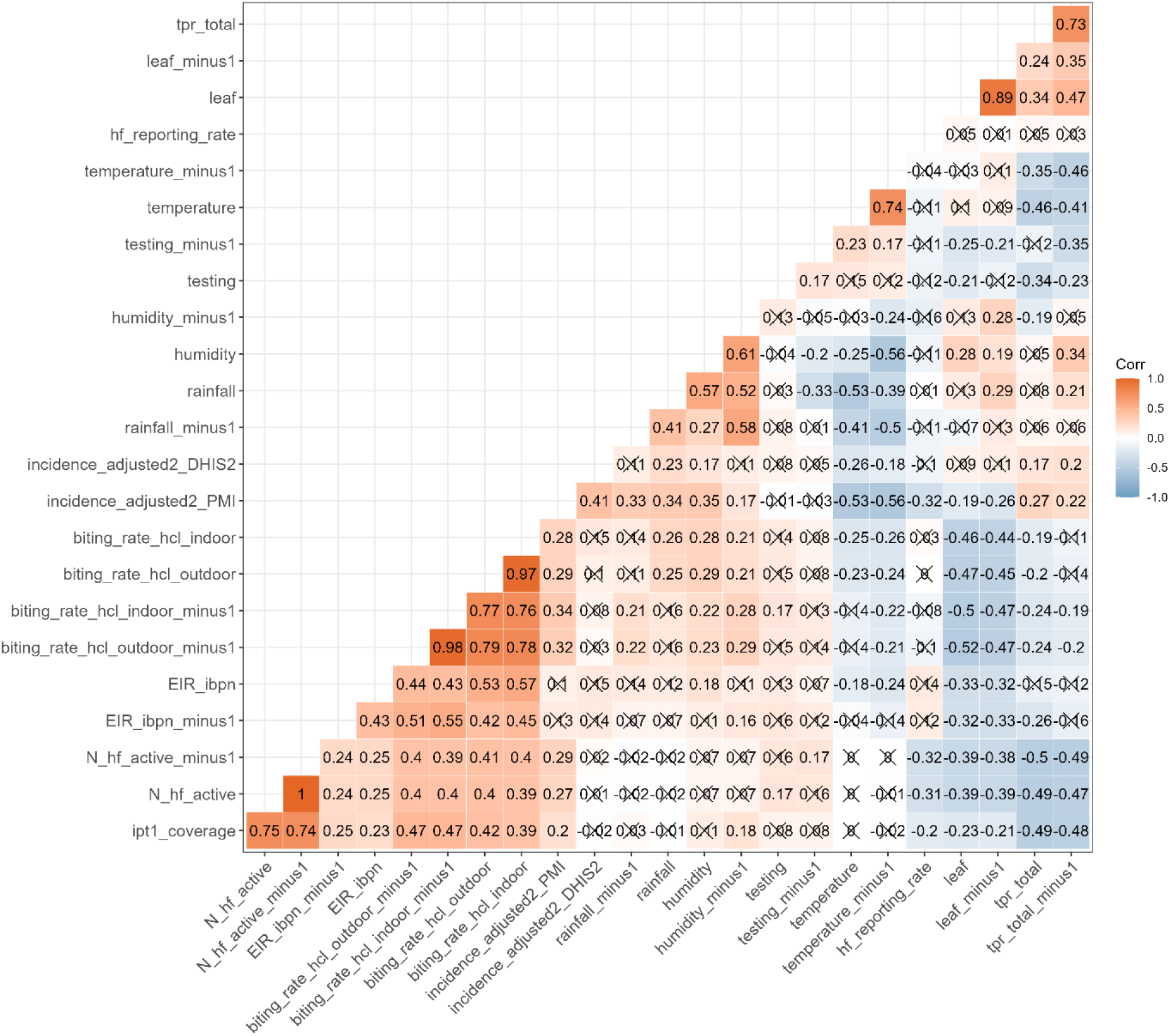
Correlation between contemporaneous variables (and their one-month lag) and incidence rates. The color gradient represents positive (red) and negative (blue) correlations, crossed-out fields are correlations that do not meet the significance criteria (Pearson p-value<0.05).

Strikingly, none of the entomological variables had significant correlations with malaria incidence data from DHIS2. On the other hand, DHIS2 incidence data significantly correlated with rainfall and humidity, albeit with smaller coefficients than PMI data. A similar pattern was observed for correlations between incidence and health surveillance yield and quality covariates (number of health facilities active, health facility reporting rates) which had significant correlations with PMI incidence data (0.27, and −0.32 resp.), whereas these same covariates did not correlate with DHIS2 incidence data. Only test positivity rates positively correlated with incidence data from both sources.

### Quantification of causal impact of IRS towards malaria incidence

Based on our findings from the correlation analysis, we included all except the entomological covariates into our Bayesian structural time series analysis (see Methods) as these were causally dependent on the intervention. We trained a model on incidence data prior to the intervention including contemporaneous variables and utilized the resulting model estimates to predict a counterfactual for the Nassian and Sakassou district during the intervention period. Finally, to measure impact, we compared the counterfactual incidence with actually recorded incidence data (see Supplementary Figure 2). Using the incidence in the IRS districts alone during the model training (Figure 3, control=none), we obtained an estimated average decrease in incidence during the first year post intervention of 30% (95%CI: 11-43%) for Nassian and 36% (95%CI: 20-48%) for Sakassou using DHIS2 data. Incidence decrease according to PMI data was not significant. Including incidence data from the control districts (Dabakala for Nassian and Beoumi for Sakassou) to the pre-intervention model training resulted in significant impact of 12% (95%CI: 3-19%) resp. 19% (95%CI: 10-27%) for PMI data and 30% (95%CI: 19-38%) resp. 43% (95%CI: 34-50%) for DHIS2 data.

**Figure 3:**
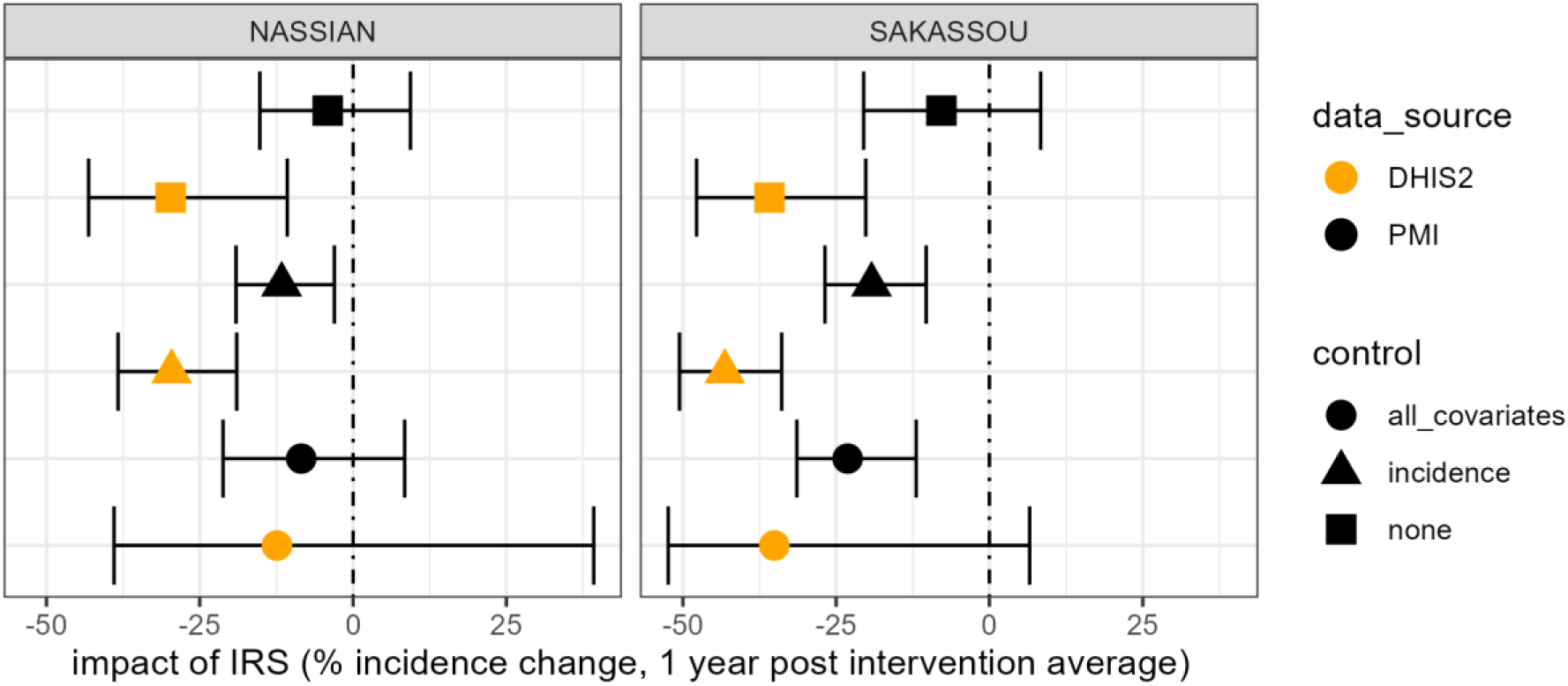
Causal impact analysis for the 2020 IRS campaign in Nassian and Sakassou. The points show the mean of the posterior distribution, the error bars represent the 95% credence interval of the posterior of the average change in incidence during 1 year post intervention. Colors distinguish data sources, shapes refer to whether only the incidence rate time series itself has been used (none), or incidence of the respective control districts (incidence) or in addition other contemporaneous variables (all_covariates). For the latter, we excluded entomological covariates (EIR_ibnp, biting_rate_hcl_outdoor, biting_rate_hcl_indoor), as these were not independent of the intervention and thus not suitable as controls.

Considering all covariates besides those related to entomology yielded an estimated impact for PMI data of 9% (not significant) for Nassian and 23% (95%CI: 12-31%) for Sakassou. No significant impact for DHIS2 data was determined in that case. The large error estimates from the credence intervals suggest a high variability in posterior variable selection.

### Contemporaneous covariates influencing the counterfactual

Utilizing the Bayesian structural time series model, we obtained regression coefficients for each of the contemporaneous covariates. Posterior distributions from the Bayesian updating stem from spike and slab priors (see Methods), such that not all coefficients were necessarily selected in each model run. We focused on the marginalized regression coefficients that consistently either had negative or positive values for 95% of the model runs in order to quantify covariates influencing the counterfactual, and consequently the evaluated impact.

E.g. a consistently positive regression coefficients would mean increased incidence for the counterfactual for an increase in the covariates, such that the overall intervention impact would be have been overestimated without the covariate, and vice versa for negative regression coefficients.

Considering empirical distributions of posterior regression coefficients (Figure 4), we observed that incidence from the control district had positive estimates across datasets and health districts with high probability of inclusion for the model selection process. Health facility reporting rates were consistently associated with negative coefficients for PMI data, suggesting that for periods with increased health facility reporting during the intervention, one would have estimated a decrease in the counterfactual and consequently a decreased impact of IRS. For DHIS2 on the other hand, test positivity rate, number of health facilities active and rainfall had positive coefficients, such that an increase in these factors might be the main driver of the overestimated impact of IRS (Figure 3). We stress that the posterior distribution of covariate regression coefficients used for the impact quantification are high-dimensional and not independent from each other such that the underlying causality might rather be a combination of many factors than just those mentioned above.

**Figure 4:**
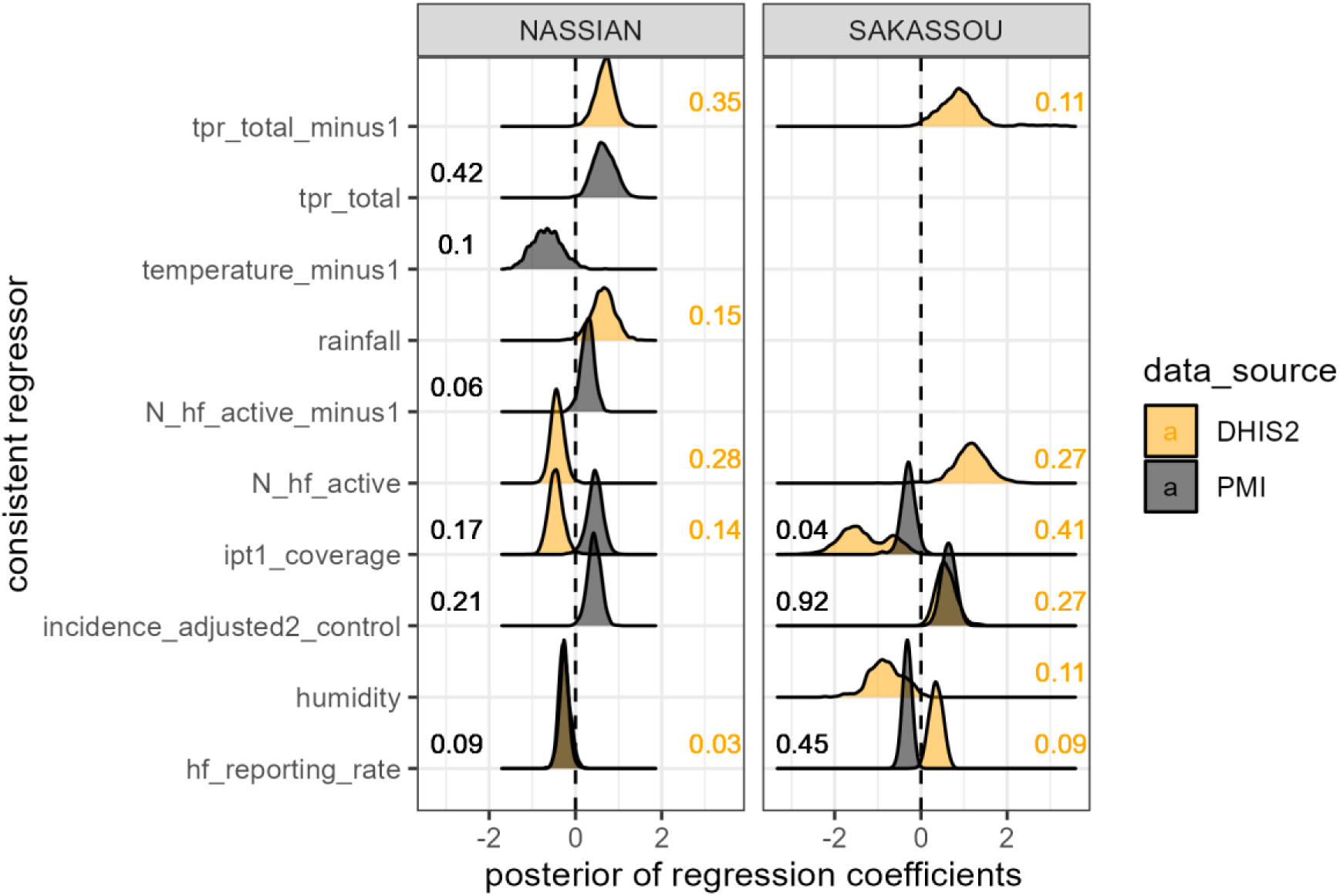
Posterior distribution of regression coefficients (with possible one-month lags) from the Bayesian causal impact analysis that have 95% of the mass either above or below zero. Positive coefficients indicate an increase in counterfactual incidence for a unit increase of the regressor, thus an overestimation of intervention impact if the regressor were not taken into account. The numbers in the graph are inclusion probabilities in the model over 10000 runs following the spike and slab prior.

### Quantification of causal impact of IRS towards entomological inoculation rates

Utilizing the same approach as for the causal impact analysis towards incidence, we also investigated causal impact towards EIR. This time we excluded malaria incidence as covariates, as these were not independent from EIR rates. We observed a strong and statistically significant causal impact of IRS on the decrease of EIR at the order of 70% averaged during one year post intervention (Figure 5). Among consistent regressors, only EIR values from control districts were chosen with high inclusion probability (Figure not shown).

**Figure 5:**
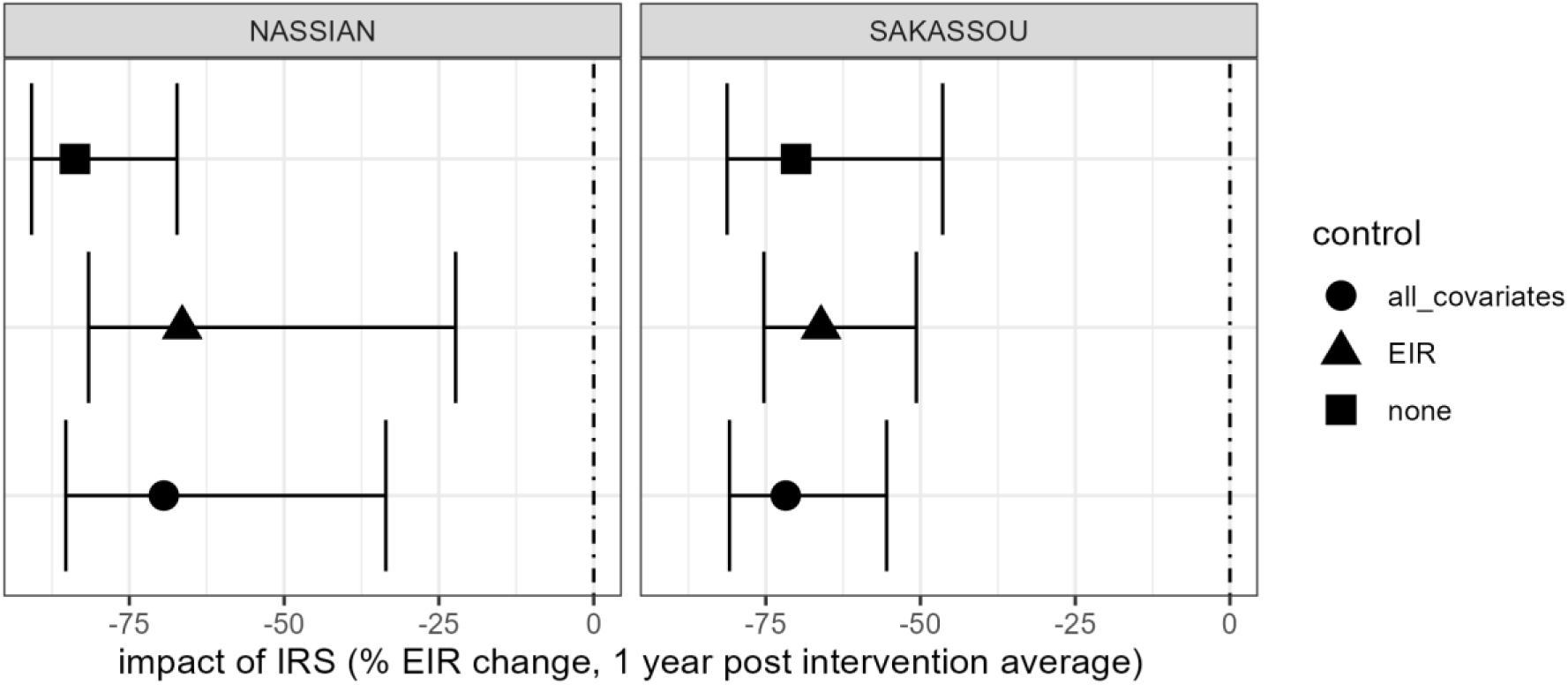
Causal impact analysis for the 2020 IRS campaign in Nassian and Sakassou. The points show the mean of the posterior distribution, the error bars represent the 95% credence interval of the posterior of the average change in entomological inoculation rate during 1 year post intervention. The control level “all covariates” in the plot refers to using all covariates in the intervention districts Nassian and Sakassou (except malaria incidence, see methods), the level “EIR” refers to using EIR values from control districts Beoumi and Dabakala in addition to EIR values in the intervention districts. The level “none” refers to using EIR data only from the intervention districts Nassian and Sakassou.

## Discussion

With improved quality and availability of routine surveillance data for malaria, retrospective impact analysis becomes an important tool to bridge the gap from efficacy in clinical trials to effectiveness at operationally relevant scales such as health districts (15). For NMCPs, retrospective impact quantification is highly relevant as it provides context-specific information to evaluate program efficiency and to plan future investments. In the present study, we interrogate clinical incidence data extracted for the first time in its full breadth from the DHIS2 routine surveillance system in Côte d’Ivoire (8). Established since 2015, this system has since seen major improvements (e.g. including new indicators, capturing community health workers and private sector) but is still underused to provide analytics for decision support. Côte d’Ivoire has rolled out a pilot of indoor residual spraying in 2020 and 2021 in two health districts thanks to support from PMI. Concurrently with this intervention, PMI has also collaborated with the NMCP to re-digitize health facility data from paper registries to quantify the impact of IRS. Our study specifically focuses on quantifying how data quality impedes impact quantification and how a Bayesian framework could inform causality between covariates and impact indicators. We summarize and contextualize our four main findings.

First, correlation analysis with remote-sensed temporal covariates such as rainfall or biting rates revealed that DHIS2 data is not following expected patterns of seasonality whereas PMI data does. Likewise, we observed highly significant correlation of PMI incidence with indicators of surveillance quality such as reporting rates and number of active health facilities, whereas DHIS2 does not. We conclude that DHIS2 data very likely does not faithfully represent incidence dynamics at monthly scales and that coarser temporal aggregation would be better suited for analysis with such data. IRS deployment and test positivity rates are the factors most strongly associated with decreasing gaps between PMI and DHIS2 incidence data across several statistical models. We suggest that viability and sensitivity of tests and their reporting should be further improved in line with observations from key-informant interviews (8).

Second, although using a completely different method, our results of 12% resp. 19% incidence decrease one year post intervention for PMI data are in good agreement with a recently published analysis (9), which reports 16% resp. 15.8% reduction in Nassian resp. Sakassou. Using DHIS2 data, our statistical models estimated much higher impact of 30% reps. 42%, which is close to 44% and 47% DHIS2-based estimates for IRS in Ghana and in Uganda (27,28). This further highlights the robustness of our results and the importance of the data source.

Third, following the Bayesian model averaging approach, test positivity rates, incidence from control districts and reporting rates of health facilities were identified as consistent regressors. Increased test positivity and incidence in control districts during the intervention period would increase counterfactual incidence and thus not taking these covariates into account would overestimate the impact of IRS. Likewise, increased facility reporting rates would decrease counterfactual incidence and not considering these would lead to underestimated impact.

Fourth, a separate analysis focused on EIR data, which to our knowledge has only been used to choose insecticides (29) but not for IRS impact quantification before. Our result of a strong causal impact of IRS of 70% decrease corroborates estimations of more than 80% vector mortality on walls published in end-of-spray reports (30,31). Transmission model studies (32) suggest that a 90% decrease in EIR in high endemic areas such as Nassian and Sakassou would yield a 38% decrease in clinical incidence under active case detection. Although causality between EIR and incidence remains elusive, our results indicate that entomological surveillance data could be used as surrogates for intervention impact.

This study has several limitations. The biggest hurdle to generalize our findings is data quality. We have extracted routine surveillance data from the DHIS2 database with the help of the monitoring and evaluation division at the NMCP. Despite upfront database validation work by the NMCP and data post-processing, our correlation analysis showed that temporal inconsistencies in the malaria incidence data remain and hamper fine-tuned time series analysis. The improvement in the PMI database concerned mainly electronic data entry, and our comparative analysis clearly points towards improved data quality, e.g. in terms of expected patterns of seasonality.

Although our Bayesian analysis considered many contemporaneous covariates, the underlying causality graph remains elementary. We focused on the relationship between each covariate and the incidence outcome and did not model the putative causal relationships between covariates.

Incidence time series from the control district were consistently identified as key to achieve high accuracy for the counterfactual model training in the intervention district. This finding suggests that incidence measurements from control districts are vital for counterfactual model parameter learning as they encode unobserved variables and processes. Relying solely on surrogate data that is independent from the intervention (e.g. rainfall, vegetation index) but does not contain incidence measurements would be insufficient to construct counterfactuals that de-convolute causal impact on malaria transmission.

From the study limitations arise also several opportunities to improve the operability of routine data for both evaluation and strategic planning. Routine correlation analysis with remote-sensed data and outlier analysis could help single out spurious signals and guide the deployment of data quality improvement measures akin to those described in PMI’s re-digitization protocol (9). Overestimating the impact of particular interventions comes to additional expenditures, which could be invested in strengthening the routine surveillance system at the level of data entry. Climate-related changes in patterns of vector dynamics and clinical malaria incidence call for systematic use of remote-sensed data and summary statistics of extreme events as contemporaneous variables to more faithfully estimate intervention impact. The main open question that arises from our study is the problem of causality within the complex dynamic process of disease transmission of malaria. Ideally, a causal diagram would single out known mechanism, such as the impact of rainfall on the infectious vector population, which should be causally related to disease incidence in the following month. Learning causal diagrams from high-quality data and using more complex observational models (e.g. over-dispersion and zero-inflation) could further improve the Bayesian inference.

In conclusion, we highlight that investments to improve data quality of routine surveillance by focusing on indicators such as test positivity and health facility reporting are key to avoiding overly optimistic estimations of the impact of IRS on clinical incidence in areas with high malaria prevalence such as Sakassou and Nassian. The careful choice of control districts for such analyses is crucial in order to factor out potential intervention confounders.

## Data Availability

All data produced in the present study are available upon reasonable request to the authors.

## Acknowledgements

We acknowledge the Global Fund, especially Paula Hacopian and Maria Walusimbi, for initializing the retrospective analysis based on DHIS2 malaria surveillance data. We thank Sarah Burnett from PMI for sharing previously published data produced through the collaboration between PATH and the National Malaria Control Program of Côte d’Ivoire. This work has been funded through the service contract N°UCP-FM/2022/347 from the Ministère de la Santé, de l’Hygiène Publique et de la Couverture Maladie Universelle de Côte d’Ivoire.

## List of abbreviations

PMI: President’s Malaria Initiative
NMCP: National Malaria Control Program
DHIS2: District health information system
DHS: Demographic and Health Survey
ITN: Insecticide-treated nets
IRS: Indoor residual spraying
RDT: Rapid diagnostic test
EIR: Entomological inoculation rate
RCT: randomized control trial
IPT1: Intermittent preventive therapy, first dose

## Supplementary Material

**Supplementary Figure 1:**
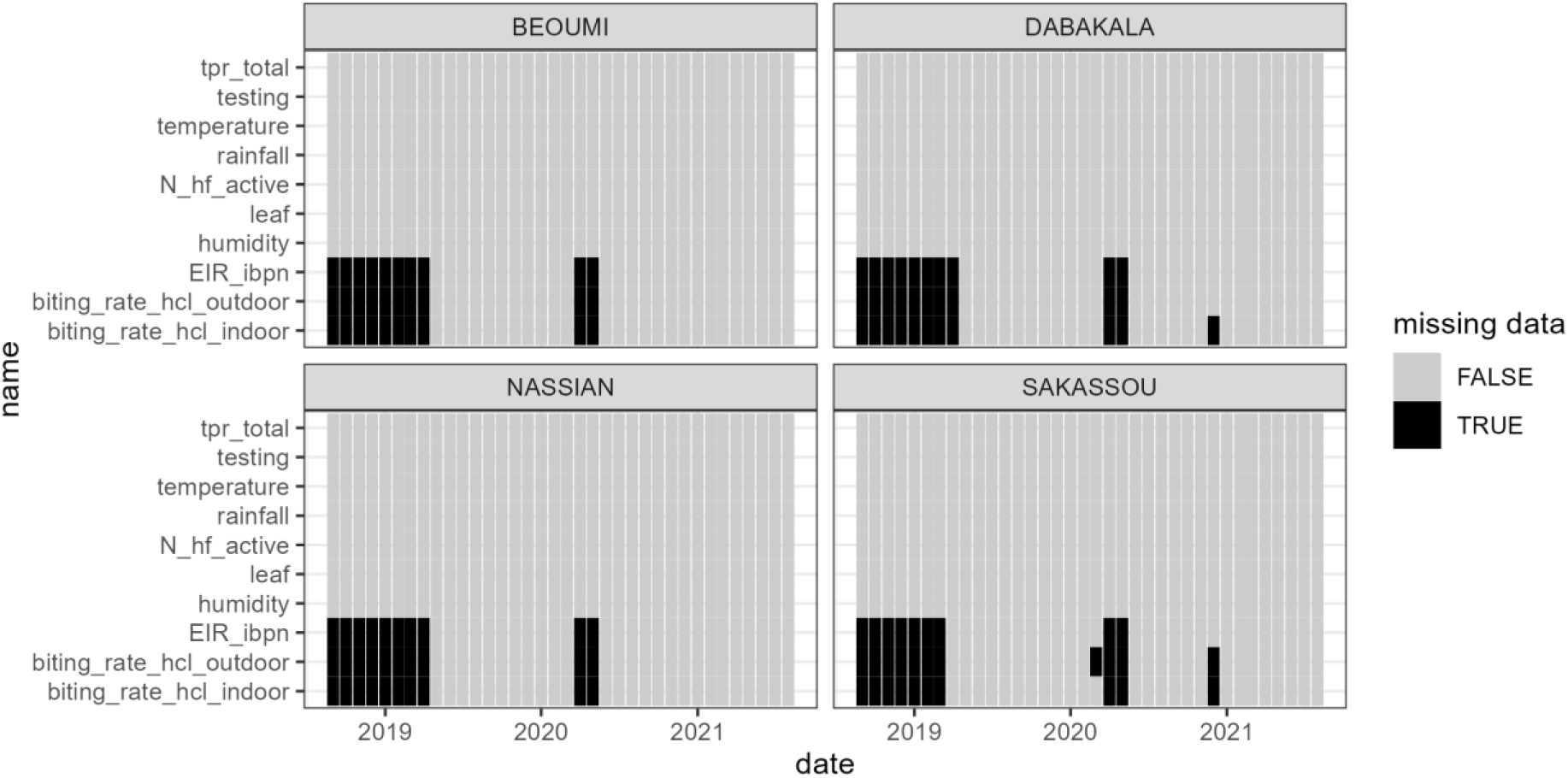
Missing data of contemporaneous covariates in the four districts of the study.

**Supplementary Figure 2:**
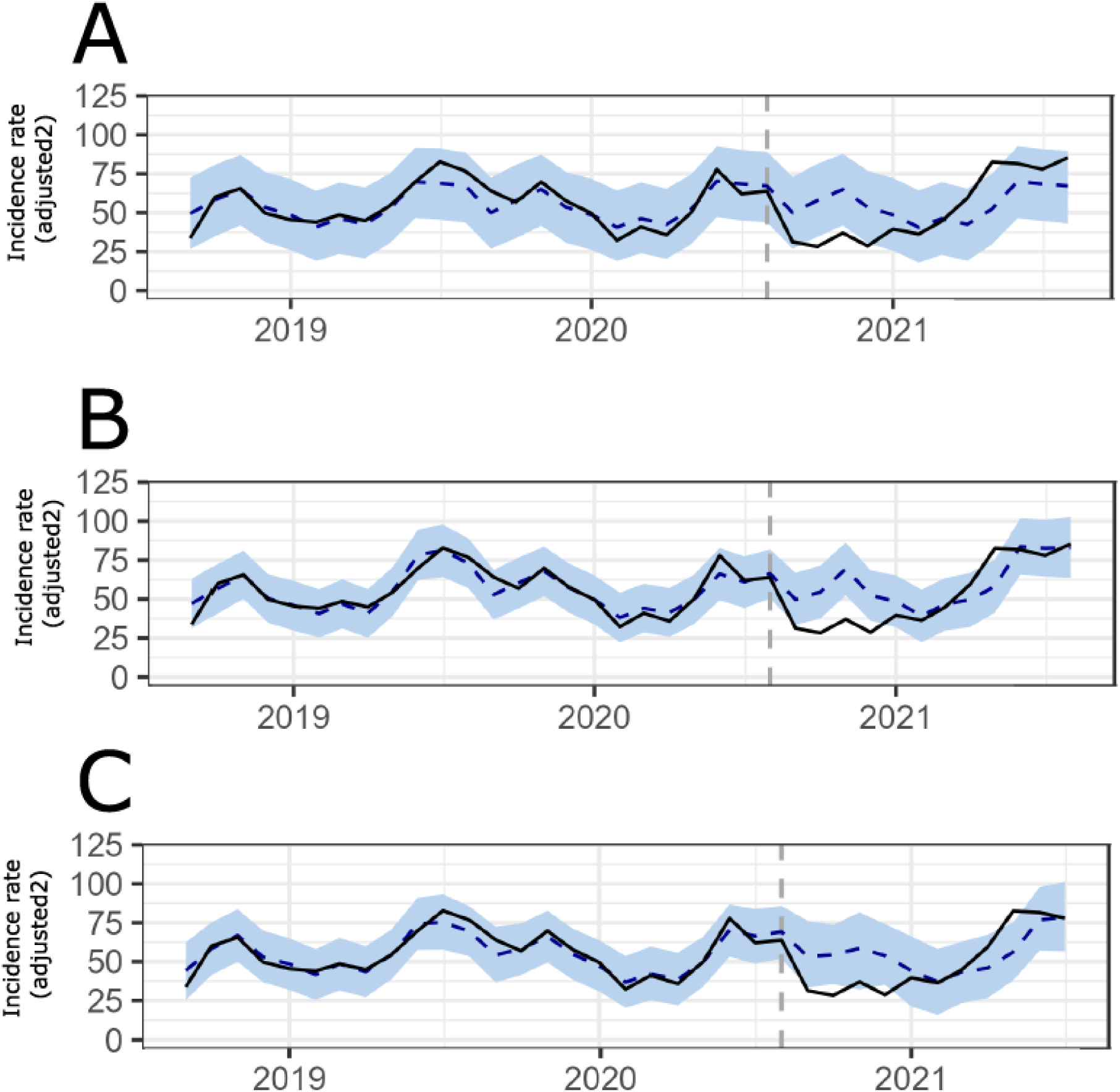
Sample run of Bayesian Structural Time Series for Nassian district with PMI data. The panels shows the incidence data (solid lines) vs. BSTS model predictions (dashed lines), before and after the start of the intervention (grey dashed line). Impact is quantified as the average % decrease in incidence after the start of the intervention comparing the recorded data (solid line) to the modeled counterfactual (dashed line). The model in panel A uses only incidence data from Nassian. In panel B, incidence data from both Nassian and the respective control district Dabakala are used. The model in panel C contains incidence data from Nassian, Dabakala and all contemporaneous covariates (see Methods section).

**Supplementary Table 1:** Description of covariates used in the causal impact analysis by source and periodicity.

| Variable name | Description | Source | monthly | yearly |
| --- | --- | --- | --- | --- |
| Rainfall | Total precipitation in m | ERA5 | X |  |
| Temperature | 2m temperature in degree Celsius | ERA5 | X |  |
| Leaf | Leaf area index, low vegetation | ERA5 | X |  |
| Humidity | 2m dewpoint temperature in degree Celsius | ERA5 | X |  |
| biting_rate_hcl_indoor | Indoor biting rate of An. gambiae and funestus combined | PMI-Vectorlink | X |  |
| biting_rate_hcl_outdoor | Outdoor biting rate of An. gambiae and funestus combined | PMI-Vectorlink | X |  |
| EIR_bpn | Infectious bites per night per person | PMI-Vectorlink | X |  |
| N_hf_active | Number of health facilities reporting cases during a month per 1000 population | DHIS2 | X |  |
| hf_reporting_rate | Number of consecutive months reporting cases per months from first report until end of year | DHIS2 |  | X |
| Testing | Number of tested (RDT or blood spot) per suspected cases | DHIS2 | X |  |
| tpr_total | Number of positive tested per total number of tested (RDT or blood spot) | DHIS2 | X |  |
| ipt1_coverage | Number of IPT doses administered during first trimester ANC visit per total number of first trimester ANC visits | DHIS2 |  | X |

**Supplementary Table 2:** Adjustment of incidence rate for presumed (i.e. untested) cases, reporting rate and health care seeking applied to both DHIS2 and PMI crude incidence.

| Adjustment | Description | Details | Formula | Data sources |
| --- | --- | --- | --- | --- |
| 0 | crude incidence | uncomplicated & complicated cases | $N_0$ | DHIS2, PMI |
| 1 | presumed (**pr**) & test positivity rate (**tpr**) | Add cases that have not been confirmed (i.e. presumed) by assuming that their chance to test positive is as **tpr** | $N_1 = N_0 + pr * tpr$ | DHIS2 |
| 2 | reporting rate (**rr**) | Take into account the annual reporting rate | $N_2 = N_1 / rr$ | DHIS2 |
| 3 | health care seeking (**s**) | Add to incidence those cases with odds of having not seeked care $(1-s)/s$ | $N_3 = N_2 + N_2 * (1-s)/s$ | DHS 2021 |

**Supplementary Table 3:** Model selection of log-linear mixed-effect models of the difference between DHIS2 and PMI incidence using backward step-wise reduction on *model1* (health system covariates only)

| regression coefficient | eliminated at step... | p-value (F test) |
| --- | --- | --- |
| IRS:hf_reporting_rate | 1 | 0.969484 |
| IRS:N_hf_active | 2 | 0.940614 |
| N_hf_active | 3 | 0.847019 |
| IRS:tpr_total | 4 | 0.758068 |
| IRS:ipt1_coverage | 5 | 0.616031 |
| IRS:testing | 6 | 0.599217 |
| testing | 7 | 0.360217 |
| hf_reporting_rate | 8 | 0.056271 |
| ipt1_coverage | 9 | 0.180847 |
| IRS | 0 | 8.62E-07 |
| tpr_total | 0 | 0.003838 |

**Supplementary Table 4:**
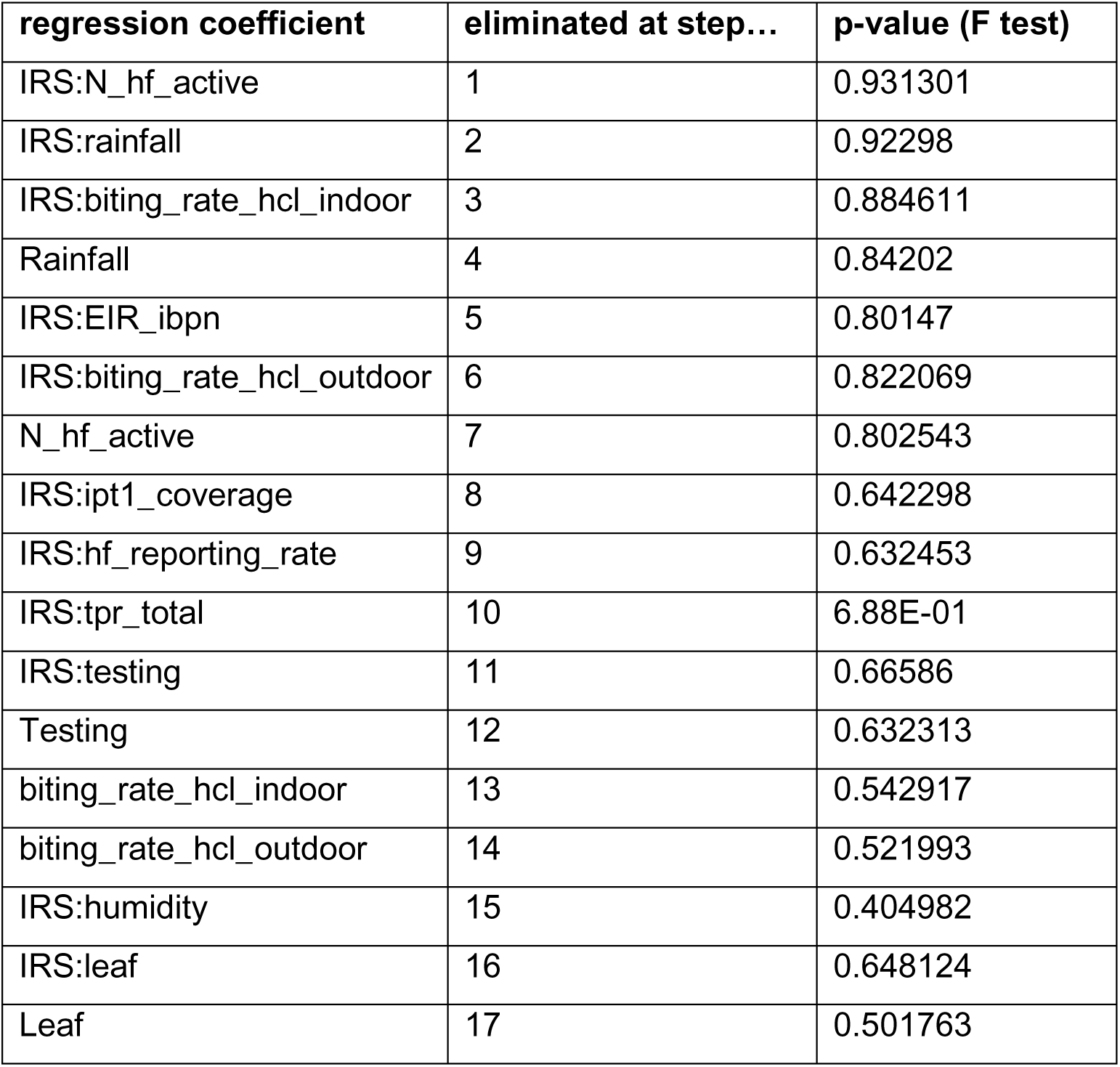
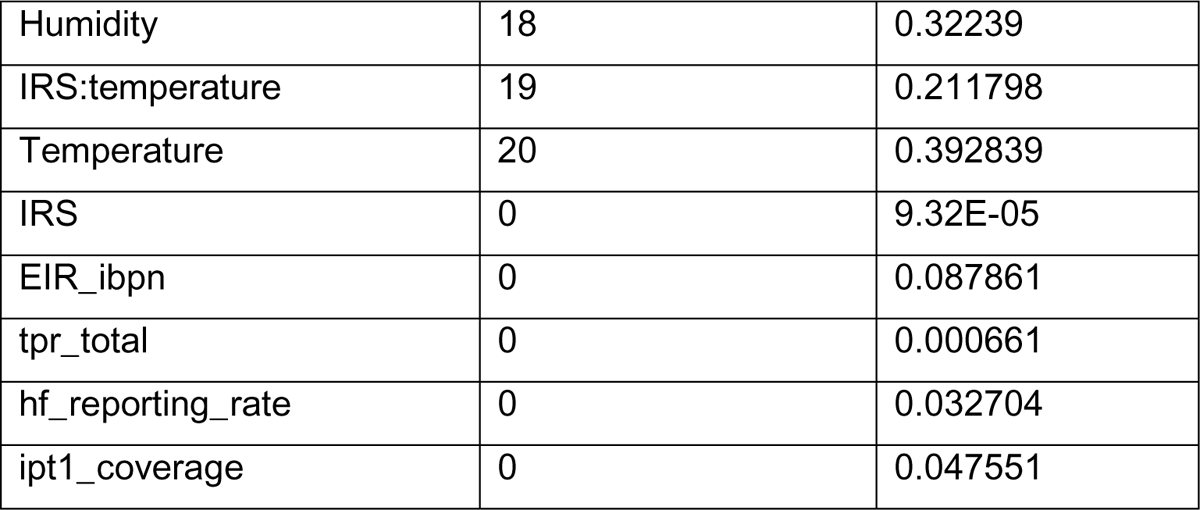
Model selection of log-linear mixed-effect models of the difference between DHIS2 and PMI incidence using backward step-wise reduction *model2* (health system covariates as well as environmental variables)

| regression coefficient | eliminated at step... | p-value (F test) |
| --- | --- | --- |
| IRS:N_hf_active | 1 | 0.931301 |
| IRS:rainfall | 2 | 0.92298 |
| IRS:biting_rate_hcl_indoor | 3 | 0.884611 |
| Rainfall | 4 | 0.84202 |
| IRS:EIR_ibpn | 5 | 0.80147 |
| IRS:biting_rate_hcl_outdoor | 6 | 0.822069 |
| N_hf_active | 7 | 0.802543 |
| IRS:ipt1_coverage | 8 | 0.642298 |
| IRS:hf_reporting_rate | 9 | 0.632453 |
| IRS:tpr_total | 10 | 6.88E-01 |
| IRS:testing | 11 | 0.66586 |
| Testing | 12 | 0.632313 |
| biting_rate_hcl_indoor | 13 | 0.542917 |
| biting_rate_hcl_outdoor | 14 | 0.521993 |
| IRS:humidity | 15 | 0.404982 |
| IRS:leaf | 16 | 0.648124 |
| Leaf | 17 | 0.501763 |
| Humidity | 18 | 0.32239 |
| IRS:temperature | 19 | 0.211798 |
| Temperature | 20 | 0.392839 |
| IRS | 0 | 9.32E-05 |
| EIR_ibpn | 0 | 0.087861 |
| tpr_total | 0 | 0.000661 |
| hf_reporting_rate | 0 | 0.032704 |
| ipt1_coverage | 0 | 0.047551 |

## References

1. Global Malaria Program WHO Geneva. World Malaria Report 2023. World Health Organization;

2. Programme National de Lutte contre le Paludisme - PNLP. Plan Stratégique National de Plaidoyer en Matière de la Lutte contre le Paludisme en Côte d’Ivoire 2018-2023. Johns Hopkins University; 2019.

3. Bhatt S, Weiss DJ, Cameron E, Bisanzio D, Mappin B, Dalrymple U, et al. The effect of malaria control on Plasmodium falciparum in Africa between 2000 and 2015. Nature. 2015 Oct 1;526(7572):207–11.

4. Pryce J, Medley N, Choi L. Indoor residual spraying for preventing malaria in communities using insecticide-treated nets - Pryce, J - 2022 | Cochrane Library. [cited 2024 Sep 9]; Available from: https://www.cochranelibrary.com/cdsr/doi/10.1002/14651858.CD012688.pub3/full

5. Hilton ER, Rabeherisoa S, Ramandimbiarijaona H, Rajaratnam J, Belemvire A, Kapesa L, et al. Using routine health data to evaluate the impact of indoor residual spraying on malaria transmission in Madagascar. BMJ Glob Health. 2023 Jul;8(7):e010818.

6. Chanda E, Coleman M, Kleinschmidt I, Hemingway J, Hamainza B, Masaninga F, et al. Impact assessment of malaria vector control using routine surveillance data in Zambia: implications for monitoring and evaluation. Malaria Journal. 2012 Dec 29;11(1):437.

7. Epstein A, Namuganga JF, Nabende I, Kamya EV, Kamya MR, Dorsey G, et al. Mapping malaria incidence using routine health facility surveillance data in Uganda. BMJ Global Health. 2023 May 1;8(5):e011137.

8. Selinger C, Angoa G. Analyse rétrospective 2012-2022 des activités de lutte contre le paludisme en Côte d’Ivoire. unpublished report; 2023.

9. Hilton ER, Gning-Cisse N, Assi A, Eyakou M, Koffi J, Gnakou B, et al. Reduction of malaria case incidence following the introduction of clothianidin-based indoor residual spraying in previously unsprayed districts: an observational analysis using health facility register data from Côte d’Ivoire, 2018–2022. BMJ Glob Health. 2024 Mar;9(3):e013324.

10. Bernal JL, Cummins S, Gasparrini A. Interrupted time series regression for the evaluation of public health interventions: a tutorial. Int J Epidemiol. 2017 Feb;46(1):348–55.

11. Turner SL, Forbes AB, Karahalios A, Taljaard M, McKenzie JE. Evaluation of statistical methods used in the analysis of interrupted time series studies: a simulation study. BMC Medical Research Methodology. 2021 Aug 28;21(1):181.

12. Gianacas C, Liu B, Kirk M, Tanna GLD, Belcher J, Blogg S, et al. Bayesian structural time series, an alternative to interrupted time series in the right circumstances. Journal of Clinical Epidemiology. 2023 Nov 1;163:102–10.

13. Liu J, Spakowicz DJ, Ash GI, Hoyd R, Ahluwalia R, Zhang A, et al. Bayesian structural time series for biomedical sensor data: A flexible modeling framework for evaluating interventions. PLoS Computational Biology [Internet]. 2021 Aug [cited 2024 Sep 9];17(8). Available from: https://www.ncbi.nlm.nih.gov/pmc/articles/PMC8412351/

14. Midelet A, Bailly S, Borel JC, Hy RL, Schaeffer MC, Baillieul S, et al. Bayesian Structural Time Series With Synthetic Controls for Evaluating the Impact of Mask Changes in Residual Apnea-Hypopnea Index Telemonitoring Data. IEEE Journal of Biomedical and Health Informatics. 2022 Oct;26(10):5213–22.

15. Burnett SM, Davis KM, Assefa G, Gogue C, Hinneh LD, Littrell M, et al. Process and Methodological Considerations for Observational Analyses of Vector Control Interventions in Sub-Saharan Africa Using Routine Malaria Data. 2025 Jan 7 [cited 2025 Mar 4]; Available from: https://www.ajtmh.org/view/journals/tpmd/112/1_Suppl/article-p17.xml

16. Hersbach H, Bell B, Berrisford P, Hirahara S, Horányi A, Muñoz-Sabater J, et al. The ERA5 global reanalysis. Quart J Royal Meteoro Soc. 2020 Jul;146(730):1999–2049.

17. The PMI VectorLink Project. The PMI VectorLink Côte d’Ivoire 2019 Annual Entomological Report, January 2019–December 2019. 2020.

18. The PMI VectorLink Project. The PMI VectorLink Côte d’Ivoire 2020 Annual Entomological Report, January 2020–December 2020. Rockville, MD. The PMI VectorLink Project, Abt Associates.; 2021.

19. The PMI VectorLink Project. The PMI VectorLink Côte d’Ivoire 2021 Annual Entomological Report, January 2021–December 2021. Rockville, MD. The PMI VectorLink Project, Abt Associates.; 2022.

20. Institut National de la Statistique - INS, Ministère du Plan et du Développement - MPD, Programme National de Lutte contre le Paludisme - PNLP, Ministère de la Santé et de l’Hygiène Publique - MSHP, ICF. République de Côte d’Ivoire Enquête de Prevalence Parasitaire du Paludisme et de l’Anémie (EPPA-CI) 2016 [Internet]. Abidjan, Côte d’Ivoire: INS/MPD/PNLP/MSHP and ICF; 2016. Available from: http://dhsprogram.com/pubs/pdf/FR330/FR330.pdf

21. Institut National de la Statistique ICF. Côte d’Ivoire Enquête Démographique et de Santé 2021 Rapport final [Internet]. Rockville, Maryland, USA et la Côte d’Ivoire: INS et ICF; 2023. Available from: https://www.dhsprogram.com/pubs/pdf/FR385/FR385.pdf

22. Honaker J, King G, Blackwell M. Amelia II: A Program for Missing Data. J Stat Soft. 2011 Dec 12;45(7):1–47.

23. Bates D, Mächler M, Bolker B, Walker S. Fitting Linear Mixed-Effects Models Using lme4. J Stat Soft. 2015 Oct 7;67(1):1–48.

24. Kuznetsova A, Brockhoff PB, Christensen RHB. lmerTest Package: Tests in Linear Mixed Effects Models. J Stat Soft. 2017 Dec 6;82(13):1–26.

25. Scott SL, Varian HR. 4. Bayesian Variable Selection for Nowcasting Economic Time Series [Internet]. Goldfarb A, Greenstein SM, Tucker CE, editors. Economic Analysis of the Digital Economy. Chicago: University of Chicago Press; 2015. p. 119–36. Available from: 10.7208/9780226206981-007

26. Brodersen KH, Gallusser F, Koehler J, Remy N, Scott SL. INFERRING CAUSAL IMPACT USING BAYESIAN STRUCTURAL TIME-SERIES MODELS. The Annals of Applied Statistics. 2015;9(1):247–74.

27. Gogue C, Wagman J, Tynuv K, Saibu A, Yihdego Y, Malm K, et al. An observational analysis of the impact of indoor residual spraying in Northern, Upper East, and Upper West Regions of Ghana: 2014 through 2017. Malaria Journal. 2020 Jul 11;19(1):242.

28. Ronald M, Humphrey W, Adoke Y, Jean-Pierre VG. Impact of population based indoor residual spraying in combination with mass drug administration on malaria incidence and test positivity in a high transmission setting in north eastern Uganda. Malaria Journal. 2023 Dec 13;22(1):378.

29. Kouassi BL, Edi C, Ouattara AF, Ekra AK, Bellai LG, Gouaméné J, et al. Entomological monitoring data driving decision-making for appropriate and sustainable malaria vector control in Côte d’Ivoire. Malar J. 2023 Jan 12;22(1):14.

30. The PMI VectorLink Project. The PMI VectorLink Côte d’Ivoire 2020 End-of-Spray Report: August 10–September 12, 2020. Rockville, MD. The PMI VectorLink Project, Abt Associates Inc.;

31. The PMI VectorLink Project. The PMI VectorLink Côte d’Ivoire 2021 End-of-Spray Report August 2–September 4, 2021. Rockville, MD. The PMI VectorLink Project, Abt Associates.;

32. Cameron E, Battle KE, Bhatt S, Weiss DJ, Bisanzio D, Mappin B, et al. Defining the relationship between infection prevalence and clinical incidence of Plasmodium falciparum malaria. Nat Commun. 2015 Sep 8;6(1):8170.

